# Improving communications in PPE: A solution for ‘landline’ telephone communication

**DOI:** 10.1101/2022.05.20.22272787

**Authors:** Timothy J Coats, Edward Pallett, Jasdip Mangat, Emma Chung

## Abstract

**Background:** Emergency care staff wearing elastomeric respiratory PPE report difficulties in communicating by telephone. We developed and tested an affordable technological solution aimed at improving telephone call intelligibility in staff wearing PPE.

**Methods:** A novel headset was created to enable a throat microphone and bone conduction headset to be used in combination with a standard hospital “emergency alert” telephone system. Speech intelligibility of a ED staff member wearing PPE was compared between the proposed headset and current practice by simultaneously recording a short-version of the Modified Rhyme Test and a Key Sentences Test. Recordings were played back to a group of blinded ED staff listening to pairs of recordings under identical conditions. The proportion of correctly identified words was compared using a paired t-test.

**Results:** Fifteen ED staff correctly identified a mean of 73% (SD 9%) words for speech communicated via the throat microphone system, compared to only 43% (SD 11%) of words for standard practice (paired t-test, p<0.001).

**Conclusions:** Introduction of a suitable headset can significantly improve speech intelligibility during ‘emergency alert’ telephone calls.

What is already known on this subject

- Wearing an elastomeric respirator mask impairs communication.
- Elastomeric respirator masks are now in common use in emergency and critical care.

What this study adds

- Components and design for a headset to be used as an adjunct to a standard hospital emergency alert telephone system are described.
- Testing of the headset confirmed a significant improvement in the intelligibility of speech when communicating by phone while wearing respiratory PPE.

## Introduction

Re-usable elastomeric respiratory protection is now commonly worn (https://en.wikipedia.org/wiki/Elastomeric_respirator) by Emergency Department (ED), Intensive Care and Operating Theatre staff treating Covid patients, as they have lower cost than long-term use of disposable PPE, provide a better fit to the face, and reduce generation of plastic waste. However, it is increasingly recognised that elastomeric masks also impair communication^[1-6]^.

Emergency Department staff at our centre reported particular difficulty in communicating by phone with ambulance paramedics during ‘pre-arrival alerts’ for incoming critically ill patients. This is a call made from the paramedic’s mobile phone or radio to a landline telephone located in the Emergency Department’s resuscitation room. The pre-arrival alert gives information about the incident, the patient’s condition and treatment give to ensure that ED teams are adequately prepared for the arrival of an incoming critically ill patient, allowing personnel, equipment, and treatments to be pre-prepared and immediately available. Even without personal protective equipment (PPE) speech intelligibility during this important conversation can be poor due to background noise (at both ends of the call); paramedics are often talking from a moving vehicle whilst undertaking patient care and there may be poor / intermittent mobile phone reception.

We noticed that ED staff in PPE answering an emergency alert call could hear what the paramedic was saying, as the ears are not covered by the respirator, but had difficulty being understood as air conduction microphones located in the telephone’s receiver handset were poor at picking up speech through the respirator mask, even when shouting. In conversations where an elastomeric respirator was worn by both parties, communication was extremely challenging and prone to error requiring repeating of information.

Communication issues in emergency medicine have potential to lead to errors or delays in patient care resulting in worse outcomes for patients and increased stress to staff. At our centre, the ED team identified an urgent need for a technological solution aimed at overcoming difficulties encountered when answering an emergency telephone call whilst wearing respiratory PPE.

The user specifications were that the technology should: (1) improve intelligibility, (2) be available without delaying the answer of an emergency call, (3) be used by multiple members of staff without cleaning – like a telephone receiver, and (4) be easily worn around a respirator mask.

## Methods

A headset combining a low-cost bone conduction earpiece and throat microphone was made suitable for connection to the hospital’s ‘emergency phone’ using a bespoke cable designed ‘in house’. Both the bone conduction earpiece and throat microphone fit around a respirator and are sealed, so can be shared between staff and are easier to clean than other input / output options. With some simple electrical connections, these two devices can be combined and linked to the ‘headset’ socket of a landline telephone via an adapter. Electronic engineering for the connection is within the capability of most hospital clinical engineering teams and the components required can be readily obtained for less than £50 ($70USD). Details of the headset setup are freely available on GitHub^[7]^, under a Creative Commons Zero v1.0 Universal licence (meaning that it can be copied without restrictions).

To evaluate the impact of the proposed headset on communication in PPE, a member of the ED team wearing an elastomeric mask was asked to read-out 4 sets of 50 words from the Modified Rhyme Test^[8]^, and 5 phrases relevant to an Emergency Department setting in the form of a Key Sentences Test^[9]^. The speaker’s voice was picked up simultaneously using two landline hospital telephones; one connected via the proposed headset and the other through detecting speech via a standard telephone receiver. High quality recordings of the reception end of the telephone call were made by calling a VoIP (Voice over Internet Protocol) platform using the headset system and conferencing-in the standard receiver so that a single audio source (the speaker) could be captured simultaneously via both devices. Audio traffic from these two phones was captured on the edge of the VoIP platform and saved as voice recordings. The audio codecs used on the call were all standard telephony quality – 64 kbit/s - the same as that of the public switched telephone network (PSTN).

The recorded words and sentences from each pair of tests (standard vs. headset) were randomised before being replayed to 15 members of ED staff, all of whom might answer the emergency phone at work. The listeners were blinded as to which method (conventional telephone handset or novel headset) had been used to make the recording. Listening tests were conducted under identical conditions within a quiet teaching room with no added background noise. Scores for each pair of simultaneous recordings, with the conventional phone handset and the throat microphone / bone conduction headset, were compared using a paired t-test. For the Sentences Test, listeners were asked to rate the intelligibility of sentences on scale of 1-5; from poor (1/5) to excellent (5/5).

## Results

The received average volume of speech at the reception end of the telephone call made through the telephone handset was 37 dB (just above a whisper) compared with 63 dB (normal speech level) using the throat microphone system. The fifteen subjects correctly identified a mean of 43% (SD 11%) words when pronounced via the telephone receiver compared to 73% (SD 9%) using the throat microphone system (paired t-test, p<0.001). Sentences were rated as a median of “Poor” (1/5) intelligibility using the telephone receiver compared to a median of “Excellent” (5/5) using the new headset.

## Discussion

Reusable elastomeric respirators have a number of advantages over disposable PPE and are preferred by staff experienced in their use^[10]^. Using disposable (FFP3) masks in areas where patients undergo potential aerosol generating procedures becomes cumulatively more expensive over time, we estimate a cost to our ED of £75,000 (USD $105,000) per year. In contrast, equipping all staff with a £20 (USD $30) elastomeric respirator, which is likely to last for several years, would cost approximately £8000 (USD $11,000), with a small ongoing cost for an annual filter change. Disposable PPE also creates a large volume of non-recyclable waste, which is reduced by the adoption reusable technologies. These advantages mean that reusable elastomeric respirator masks are likely to remain in use in health services, despite their associated communication issues.

Each different type of disposable FFP3 mask needs to be ‘fit tested’, meaning that every member of staff needs to be re-tested at every change of manufacturer. An elastomeric mask only needs to be tested once to ensure a good fit to the wearer. With increased demand for respiratory protection, FFP3 masks can be in short supply, so the use of reusable elastomeric masks gives staff and employers confidence that appropriate protection will always be available.

Communication requirements within a hospital environment differ from those of other environments where PPE respirators may be required, such as the police, military, or fire service. Current communications solutions developed for police, army or fire service do not fulfil the needs of the clinical workplace, so novel appropriate and cost-effective solutions need to be developed to support the ongoing use of clinical PPE.

Answering an emergency phone requires a headset system that does not rely on transmission of speech through air (which is greatly impaired by elastomeric respirators), is cleanable, can be rapidly put on over a respirator mask, and that can be shared between staff. This study shows that the combination of a throat microphone and bone conduction headset fulfils these requirements, improving isolated word speech intelligibility by ∼30%. We only tested a device that was designed for a landline telephone, however many other devices are used for communication in hospital emergency care (such as mobile phone, radios and messaging systems). The impact of PPE on these other systems has not been assessed.

The electrical engineering skills needed to create an appropriate connection is within the skillset of most hospital Clinical Engineering teams. This headset is now being used as a permanent headset option on the ‘prehospital alert’ telephone in our Emergency Department. Feedback from staff has given real world confirmation that emergency communication became easier using our medical communication headset, although putting on two separate devices (the throat microphone and bone conduction headset) was described by some as “awkward” (putting on the two devices took 10 to 15 seconds, which may seem like a long time when the emergency phone is ringing). Future developments might integrate these two components into one device.

This study has a number of limitations. We have not investigated the impact of a large number of factors which may affect communication (with or without PPE) in emergency care, such as (a) pitch of voice (inc male / female difference), (b) rate of speech (anxiety in emergency situation), (c) regional accents, (d) use of first language (for example the NHS in the UK has many staff for whom English is not their first language), (e) age of clinician (volume and frequency range changes).We also only tested one type of elastomieric mask, when there are a huge range fo designs on teh market. Future work could explore differences in intelligibility between different speakers and masks. A further limitation was that our recordings and tests were conducted in a quiet environment (which is the standard for intelligibility testing). However this is not representative of an Emergency Department environment, and the presence of background noise is likely to reduce speech intelligibility using the telephone receiver still further. Future work should explore the impact of background noise at both the pre-hospital and ED ends of the call. The tested intelligibility using the standard validated test may not be generalisable to error reduction in emergency care as the MRT features lists of single words and does not consider the brain’s capacity to make sense of sounds when presented in contextual sentences.

An elastomeric mask both attenuates and distorts speech, reducing both loudness and clarity. Our novel headset compensates for this loss of volume associated with wearing PPE. Although speech was slightly distorted due to the use of a throat microphone, this did not seem to matter, perhaps because the human brain is well adapted to extract meaning from speech. So when wearing a respirator intelligibility was higher using the novel device than when communicating using just a telephone handset. Overall, our novel headset significantly improved intelligibility and it is likely that patient safety would be improved by widespread adoption of such a device to avoid mis-communication of emergency care information between ambulance service personnel and the ED.

## Data Availability

Data is available through contact with the authors.

## Notes

### Competing Interest Statement

The authors have declared no competing interest.

### Funding Statement

This study did not receive any funding

### Author Declarations

University of Leicester. Medicine and Biological Sciences Research Ethics Committee. Ethics Reference: 28021-emlc1-ls:cardiovascularsciences Ethics approval granted 03/November/2020.

