## Appendix for "Improving communications in PPE: A solution for ‘landline’ telephone communication"

### Description of how to put on the device

The two parts of the device were stored on small hooks next to the wall mounted emergency telephone. When the phone rang the clinician walked over to it and first put the throat microphone onto their neck, locating the microphone high in the next to the larynx. The bone conduction headset were then place in front of the ears. The clinician then pressed the 'headset' button on the telephone which started the connection. At the end of the conversation the clinician pressed the 'headset' button on the telephone again to terminate the connection. A poster by the telephone reminded the clinician to take off the headset before walking away. Putting on both components of the headset took about 10 seconds.

The poster demonstrating the procedure can be found at:
https://github.com/TimCoats/telephone-headset

**Test words used in the modified Matching Rhyme Test (MRT).**

The first two sets of words (1X and 1Y) from the MRT were used (50 words in each). The MRT is designed so that the phonetics of the words matches the frequency of different sounds in spoken English:

| | Words 1X | | Words 1Y |
|---|---|---|---|
| 1 | sick | 1 | tick |
| 2 | heat | 2 | neat |
| 3 | pup | 3 | pub |
| 4 | shook | 4 | look |
| 5 | tip | 5 | lip |
| 6 | race | 6 | ray |
| 7 | gang | 7 | hang |
| 8 | kill | 8 | bill |
| 9 | math | 9 | mad |
| 10 | tale | 10 | pale |
| 11 | save | 11 | same |
| 12 | peak | 12 | peach |
| 13 | kid | 13 | kill |
| 14 | sag | 14 | sat |
| 15 | sin | 15 | sill |
| 16 | hold | 16 | cold |
| 17 | but | 17 | bug |
| 18 | lay | 18 | lame |
| 19 | gun | 19 | run |
| 20 | rust | 20 | dust |
| 21 | pack | 21 | pan |
| 22 | did | 22 | din |
| 23 | fit | 23 | hit |
| 24 | tin | 24 | fin |
| 25 | tease | 25 | teak |
| 26 | went | 26 | sent |
| 27 | sub | 27 | sung |
| 28 | red | 28 | wed |
| 29 | not | 29 | tot |
| 30 | dung | 30 | dun |
| 31 | pip | 31 | pit |
| 32 | seethe | 32 | seek |
| 33 | say | 33 | pay |
| 34 | west | 34 | rest |
| 35 | pace | 35 | pave |
| 36 | back | 36 | bath |
| 37 | pop | 37 | shop |
| 38 | big | 38 | fig |
| 39 | tam | 39 | tap |
| 40 | cane | 40 | case |
| 41 | came | 41 | game |
| 41 | came | 41 | game |
| 42 | boil | 42 | soil |
| 43 | fin | 43 | fit |
| 44 | cuff | 44 | cuss |
| 45 | feel | 45 | eel |
| 46 | mark | 46 | bark |
| 47 | heal | 47 | heap |
| 48 | den | 48 | men |
| 49 | law | 49 | saw |
| 50 | beat | 50 | beak |

In the MRT each set of words is repeated for each test condition (New Device and Control) giving 4 Word List / Answer Form combinations (100 words for each test condition). The order of the words on the answer forms is designed to minimise any learning effect, as the order of words on the answer form is different when the words are repeated:

| | Form 1X | Form 1Y |
|---|---|---|
| **Words 1X** | Control | New Device |
| **Words 1Y** | New Device | Control |

The subjects listening do not know whether they are hearing a set of words recorded using the new device or a control set (test subjects blinded to mode of recording).

Subjects were asked not to change their position during the tests, as there is a natural inclination to lean forwards when words are difficult to hear (and so potentially cause bias by decreasing the distance to the sound source).

The recordings used can be found at:
https://github.com/TimCoats/telephone-headset
WARNING: The physics of sound and audio is complex. Listening to recordings in unscientific, non-standard conditions can easily give a misleading impression (it can either over-emphasise or under-emphasise any differences).

**Answer sheets used in the modified Matching Rhyme Test**

On an answer sheet that corresponded (X or Y) to the words list being read out The subjects circled the word that they heard (within the MRT the order of words on the answer sheets are varied so any learning effect is minimised).

| **Form 1X** | | | | | | | | | | | | | | |
|---|---|---|---|---|---|---|---|---|---|---|---|---|---|---|
| lick | pick | tick | 14 | sad | sass | sag | 27 | sung | sup | sun | 40 | cave | cane | came |
| wick | sick | kick | | sat | sap | sack | | sud | sum | sub | | cape | cake | case |
| seat | meat | beat | 15 | sip | sing | sick | 28 | red | wed | shed | 41 | game | tame | name |
| heat | neat | feat | | sin | sill | sit | | bed | led | fed | | fame | same | came |
| pus | pup | pun | 16 | sold | told | hold | 29 | hot | got | not | 42 | oil | foil | toil |
| puff | puck | pub | | cold | gold | fold | | tot | lot | pot | | boil | soil | coil |
| look | hook | cook | 17 | buck | but | bun | 30 | dud | dub | dun | 43 | fin | fit | fig |
| book | took | shook | | bus | buff | bug | | dug | dung | duck | | fizz | fill | fib |
| tip | lip | rip | 18 | lake | lace | lame | 31 | pip | pit | pick | 44 | cut | cub | cuff |
| dip | sip | hip | | lane | lay | late | | pig | pill | pin | | cuss | cud | cup |
| rate | rave | raze | 19 | gun | run | nun | 32 | seem | seethe | seep | 45 | feel | eel | reel |
| race | ray | rake | | fun | sun | bun | | seen | seed | seek | | heel | peel | keel |
| bang | rang | sang | 20 | rust | dust | just | 33 | day | say | way | 46 | dark | lark | bark |
| gang | hang | fang | | must | bust | gust | | may | gay | pay | | park | mark | hark |
| hill | till | bill | 21 | pan | path | pad | 34 | rest | best | test | 47 | heap | heat | heave |
| fill | kill | will | | pass | pat | pack | | nest | vest | west | | hear | heath | heal |
| mat | man | mad | 22 | dim | dig | dill | 35 | pane | pay | pave | 48 | men | then | hen |
| mass | math | map | | did | din | dip | | pale | pace | page | | ten | pen | den |
| tale | pale | male | 23 | wit | fit | kit | 36 | bat | bad | back | 49 | raw | paw | law |
| bale | gale | sale | | bit | sit | hit | | bath | ban | bass | | saw | thaw | jaw |
| sake | sale | save | 24 | din | tin | pin | 37 | cop | top | mop | 50 | bead | beat | bean |
| same | safe | sane | | sin | win | fin | | pop | shop | hop | | beach | beam | beak |
| peat | peak | peace | 25 | teal | teach | team | 38 | fig | pig | rig | | | | |
| peas | peal | peach | | tease | teak | tear | | dig | wig | big | | | | |

**Form 1Y**

| | | | | | | | | | | | | | | |
|---|---|---|---|---|---|---|---|---|---|---|---|---|---|---|
| kick | lick | sick | 14 | sack | sad | sap | 27 | sup | sub | sud | 40 | cake | came | cave |
| tick | wick | pick | | sag | sat | sass | | sum | sun | sung | | cane | case | cape |
| neat | beat | seat | 15 | sit | sip | sill | 28 | wed | fed | bed | 41 | tame | came | fame |
| meat | feat | heat | | sick | sin | sing | | led | shed | red | | same | name | game |
| pun | puff | pup | 16 | fold | sold | gold | 29 | pot | hot | lot | 42 | toil | boil | foil |
| pub | pus | puck | | hold | cold | told | | not | tot | got | | coil | oil | soil |
| hook | shook | book | 17 | but | bug | bus | 30 | duck | dud | dung | 43 | fig | fizz | fit |
| took | cook | look | | buff | bun | buck | | dun | dug | dub | | fib | fin | fill |
| lip | hip | dip | 18 | late | lake | lay | 31 | pit | pin | pig | 44 | cuss | cud | cup |
| sip | rip | tip | | lame | lane | lace | | pill | pick | pip | | cut | cub | cuff |
| rake | rate | ray | 19 | run | bun | fun | 32 | seethe | seek | seen | 45 | heel | peel | keel |
| raze | race | rave | | sun | nun | gun | | seed | seep | seem | | feel | eel | reel |
| fang | bang | hang | 20 | dust | gust | must | 33 | say | pay | may | 46 | mark | bark | dark |
| sang | gang | rang | | bust | just | rust | | gay | way | day | | lark | hark | park |
| will | hill | kill | 21 | path | pack | pass | 34 | best | rest | nest | 47 | heath | heave | heap |
| bill | fill | till | | pat | pad | pan | | vest | test | rest | | heat | heal | hear |
| map | mat | math | 22 | dip | dim | din | 35 | page | pane | pace | 48 | then | den | ten |
| mad | mass | man | | dill | did | dig | | pave | pale | pay | | pen | hen | men |
| pale | sale | bale | 23 | fit | hit | bit | 36 | bass | bat | ban | 49 | law | saw | paw |
| gale | male | tale | | sit | kit | wit | | back | bath | bad | | jaw | raw | thaw |
| sane | sake | safe | 24 | tin | fin | sin | 37 | hop | cop | shop | 50 | beat | beak | beach |
| save | same | sale | | win | pin | din | | mop | pop | top | | beam | bean | bead |
| peak | peach | peas | 25 | tear | teal | teak | 38 | dig | wig | big | | | | |
| peal | peace | peat | | team | tease | teach | | fig | pig | rig | | | | |

**Words used in 'Sentences Test'**

*Adapted from the audio commands script used by: Schumacher J, Arlidge J, Dudley D, et al. First responder communication in CBRN environments: FIRCOM-CBRN study. Emerg Med J 2019;36(8):456-58.*

### Can you hear me?

### You are at Leicester Royal Infirmary Emergency Department!

### Are you alright?

### Do you have any pain?

### Please open your eyes!

### Squeeze my fingers

### Do you have any problems breathing?

### Please stay still while I give you this injection

### Please go into the tent and undress.

### Please step into the shower and wash everywhere using this cloth

After listening to the sentences, the subjects rated the intelligibility on a 5-point scale as "Excellent, very good, good, fair, poor".

### Results Table

| **Subject** | **Control** | **New Device** | **Control** | **New Device** | **Total Control** | **Total Device** | **Difference** |
|---|---|---|---|---|---|---|---|
| 1 | 29 | 30 | 9 | 31 | 38 | 61 | +23 |
| 2 | 21 | 36 | 16 | 37 | 37 | 73 | +36 |
| 3 | 31 | 37 | 22 | 44 | 53 | 81 | +28 |
| 4 | 23 | 35 | 17 | 34 | 40 | 69 | +29 |
| 5 | 27 | 33 | 12 | 34 | 39 | 67 | +28 |
| 6 | 28 | 34 | 15 | 44 | 43 | 78 | +35 |
| 7 | 38 | 42 | 30 | 43 | 68 | 85 | +17 |
| 8 | 33 | 43 | 30 | 45 | 63 | 88 | +25 |
| 9 | 20 | 31 | 16 | 31 | 36 | 62 | +26 |
| 10 | 26 | 31 | 18 | 34 | 44 | 65 | +21 |
| 11 | 21 | 34 | 12 | 35 | 33 | 69 | +36 |
| 12 | 16 | 29 | 10 | 32 | 26 | 61 | +35 |
| 13 | 19 | 39 | 19 | 40 | 38 | 79 | +41 |
| 14 | 28 | 38 | 16 | 37 | 44 | 75 | +31 |
| 15 | 22 | 38 | 17 | 37 | 39 | 75 | +36 |
| **Total** | 382 | 530 | 259 | 558 | 641 | 1088 | +447 |
| **Percentage correct** | 51% | 71% | 35% | 74% | 43% | 73% | |
